# Culture method surveillance of *Vibrio cholerae* in a non-sewered sanitation refugee camp setting: Dzaleka Camp, Malawi

**DOI:** 10.1101/2025.01.30.25321306

**Authors:** Brandie Banner Shackelford, Petros Chigwechokha, Lucious Ziba, Christopher Misomali, Mphatso Kanjiru, Patrick Buleya, Ruth Lusungu Nyirenda, Marlene K. Wolfe, Rochelle H. Holm

## Abstract

Refugees living in camps are particularly vulnerable to infectious disease outbreaks because of overcrowding, inadequate preventative health care, and limited water, sanitation, and hygiene services. As a result of the high disease prevalence, surveillance of cholera in Malawi provides a strong rationale for site and pathogen selection for refugee camp wastewater and environmental surveillance feasibility studies. We conducted a study in Dzaleka camp, Malawi, during a nationwide cholera outbreak. Incentivized refugee volunteers took samples for 19 weeks from seven high-use public pit latrines and a vacuum pump truck used for desludging fecal sludge. The National Microbiological Reference Laboratory of the Public Health Institute of Malawi used culture methods with confirmation via VITEK^®^ MS or Analytical Profile Index to detect *Vibrio cholerae*. Academic partners provided technical input, training, and quality assurance through the study. The results were reviewed weekly at partner-coordination meetings. No *V. cholerae* was detected in samples, but one quality assurance sample tested positive during the study period. Here, we discuss the unique challenges of conducting wastewater and environmental surveillance in refugee camp settings, including our operational framework. This study provides a framework for up-scaling wastewater and environmental surveillance efforts in other humanitarian contexts through robust partnerships, processes and in-country tools.

**Graphical Abstract:** 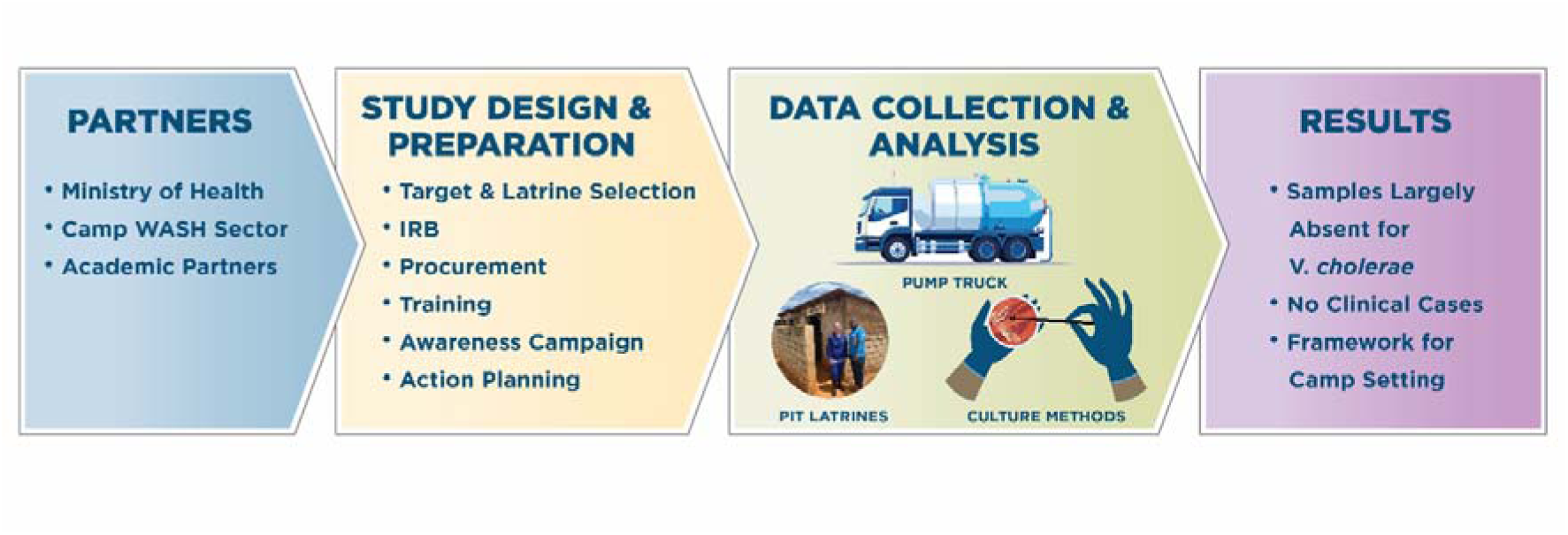

**Highlights:**

- A refugee camp in Malawi was investigated during a nationwide cholera outbreak
- Samples were collected for 19 weeks from seven pit latrines and a pump truck
- *Vibrio cholerae* was largely absent using culture methods
- Surveillance of non-sewered sanitation in a camp setting is achievable through partnerships

## 1. Introduction

Wastewater and environmental surveillance (WES) has been used across a range of geographic scales and infectious diseases, from single buildings to communities across a range of populations (Adams et al., 2024; Kilaru et al., 2023; Rondeau et al., 2023), but its application in areas with ever-increasing populations, without networked sewer systems, and especially in refugee camp settings, has been limited. Individuals affected by emergencies are particularly vulnerable to infectious diseases because of disruptions in essential water and sanitation services, high population density, and inadequate healthcare (Behnke et al., 2020; Cooper et al., 2021; Hammer et al., 2018; Jaber et al., 2024; Levy et al., 2016; Shackelford et al., 2020). Furthermore, refugees and asylum seekers may come from countries with different public health risks and arrive in host countries after experiencing inadequate transit conditions and inconsistent primary healthcare access (WHO, 2021). Health clinics in resource-limited emergency settings often lack the capacity to confirm and manage infectious disease outbreaks (Ario et al., 2022; Connolly et al. 2004), hence WES could be a useful tool for screening and surveillance to supplement clinical surveillance efforts. WES may offer an important public health surveillance tool in densely populated refugee camps with non-networked sanitation, resulting in stretched water, sanitation, and hygiene (WASH) infrastructure and gaps in healthcare.

Researchers have recently been contextualizing existing WES methodologies for low-resource settings, including creating strategies for sampling non-sewered sanitation systems (NSSS) (Capone et al., 2020; Holm et al., 2023; Jakariya et al., 2022; Keshaviah et al., 2023; Pocock et al., 2023) such as leveraging existing capacity for adapting clinical culture-based methods (Chigwechokha et al., 2024; Holm et al., 2023). Environmental surveillance for polio has been practiced for many years in resource-limited areas (Asghar et al., 2014). The potential for WES to benefit health systems in humanitarian crises was illustrated in Gaza in 2023, where polio was detected in wastewater before any clinical cases were reported (Global Polio Eradication Initiative, 2024). Although this looks practical elsewhere and for an expanded set of targets that are becoming standard in global wastewater monitoring practice, there has been a gap in implementation science examining the feasibility of starting up WES in a refugee camp setting with in-country analysis.

Malawi has not fully integrated WES into its public health system, although its potential for WES is enormous (Chigwechokha et al., 2024; Holm et al,. 2023), including the refugee settings in the country. There is high public support for WES from Malawian and Malawi refugee camp residents (Jeboda et al., 2024). Between 2022 and 2024, Malawi experienced a nationwide cholera outbreak (Chaguza et al., 2024; Republic of Malawi Ministry of Health, 2024). Due to the high disease prevalence, surveillance of cholera in Malawi provides a strong rationale for site and pathogen selection for a refugee camp feasibility study. The current study aimed to evaluate the feasibility of WES in a non-sewered sanitation refugee camp setting (Dzaleka Camp, Malawi) by sampling both high-use pit latrines and a vacuum desludging pump truck and monitoring *Vibrio cholerae* during an active outbreak. Although environmental surveillance for polio has existing guidelines in low resource contexts (WHO, 2023), the operationalizing of WES in refugee camps requires a new (non-standard) framework.

## 2. Material and methods

### 2.1 Study area

The study was conducted at Dzaleka Camp, situated in Dowa district, Malawi (Fig. 1). The United Nations High Commissioner for Refugees (UNHCR) reported that Malawi hosted 54,952 refugees and asylum seekers as of July 31, 2024, with 16.5% being children under five years old. The government mandates that refugees and asylum seekers reside within the 224 hectares camp. The Dzaleka camp is unfenced, allowing individuals from nearby host communities to access healthcare, education, and economic activities, and in some cases, to use the available sanitation facilities. The entire camp is serviced by non-sewered sanitation systems; no homes or institutions are connected to a central wastewater system.

**Fig. 1.**
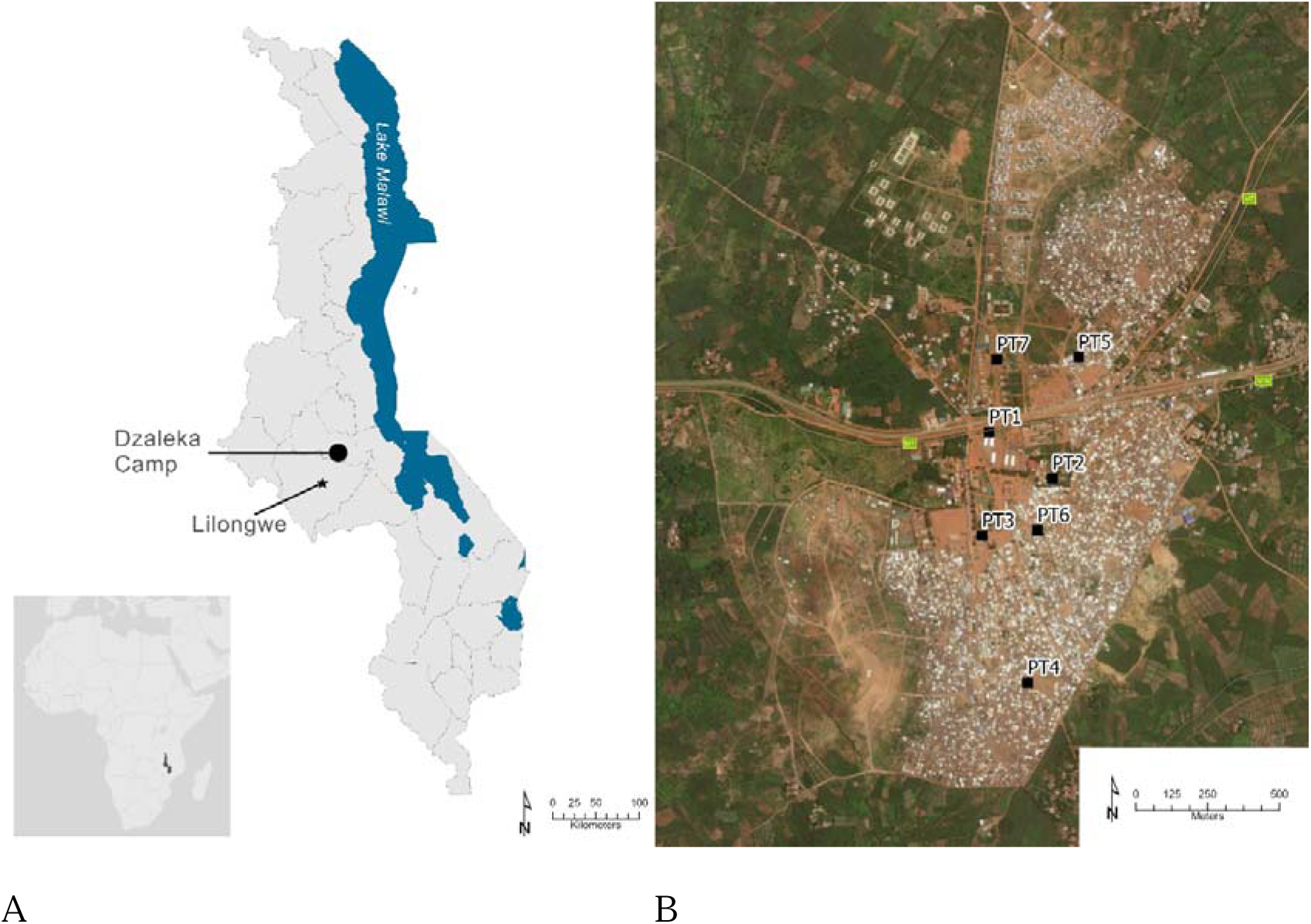
Study area, Dzaleka camp, Malawi. Panel A: The camp is located in Dowa district, approximately 40 km from the capital city Lilongwe. Panel B: Location of the seven high-use pit latrines sampled at the camp.

To determine the sampling sites, the research team first identified 65 publicly-accessible pit latrines. We then selected “high-use” pit latrines in discussions with refugee hygiene promoters. Next, an on-site feasibility check was conducted to ensure that the pit latrines had enough fecal waste for the sampling device to reach (waste was within 1 m of the ground surface) and had a roof tall enough for the sampling device to enter (>1.5 m roof height). To ensure anonymity of samples, the research team verified that the latrines supported a large number of individual contributors by recording the number of users entering the pit latrine (Supplementary Text). User counting was carried out at each site for at least 4 h on at least five different days, ensuring it occurred on distinct days of the week and times of day to account for variability (e.g., market days and church days). The number of pit latrine stances ranged from 1 to 11. If applicable, researchers also noted that if a certain pit latrine stance was more commonly used than others, sampling was prioritized for that stance (Table S1). Seven pit latrines were selected for sampling within the camp (Fig. 1).

### 2.2 Wastewater and environmental sample collection

Prior to sample collection, refugee hygiene promoters, government health surveillance assistants, and refugee community leaders informed camp residents about the project through house visits, local radio programs, and camp notice boards. Samples were collected from the seven public pit latrines (Table S2) and a vacuum desludging pump truck that emptied pit latrines of fecal waste, herein referred to as a “pump truck.” Grab samples were collected weekly on Mondays (or Tuesday this was a public holiday). Pit latrine sampling occurred from March 5, 2024, through July 9, 2024, which included parts of the rainy and cold seasons in Malawi. Pump truck sampling began on March 25, 2024, owing to delays in camp contracting (Table S3). The number of pit latrine sample sites started with four in the first week to allow sample collectors and laboratory technicians to become familiar with the procedures and increased to seven weekly sites thereafter.

Pit-latrine samples were collected by three incentive-based refugee volunteers who wore full personal protective equipment (PPE). The sampling team received an oral cholera vaccine and hands-on training before the study began. A previous study in Malawi had found that the depth of sampling in pit latrines did not impact the detection of pathogens (Capone et al., 2021). Therefore, the sampling team used a locally-fabricated sampling device: a two-meter metal pole welded to a 500 mL metal cup with a spout. The sampling team stirred the pit latrine fecal sludge three times using the sampling device to homogenize the material prior to sample collection. In instances when the fecal sludge was too thick to be sampled, the sampling team added non-chlorinated water. A rubber spatula was used to transfer the sample to a collection bottle, when necessary (Fig. 2A). A 500-mL sample was collected at each site into either sterile 250 mL glass sample bottles, 200 mL sterile plastic bottles, or sterile conical 50 mL plastic tubes. Initially, glass sample bottles were used; however, the research team switched to disposable plastic bottles because sterilizing glass was burdensome. A double volume from at least one site each week was collected for quality assurance analysis on a rotation, so that each site was captured for quality assurance over the course of the study. The sampling device and spatula were thoroughly washed with water and sterilized with 70% ethanol between samples. The sampling team donned new gloves after each sample.

**Fig. 2.**
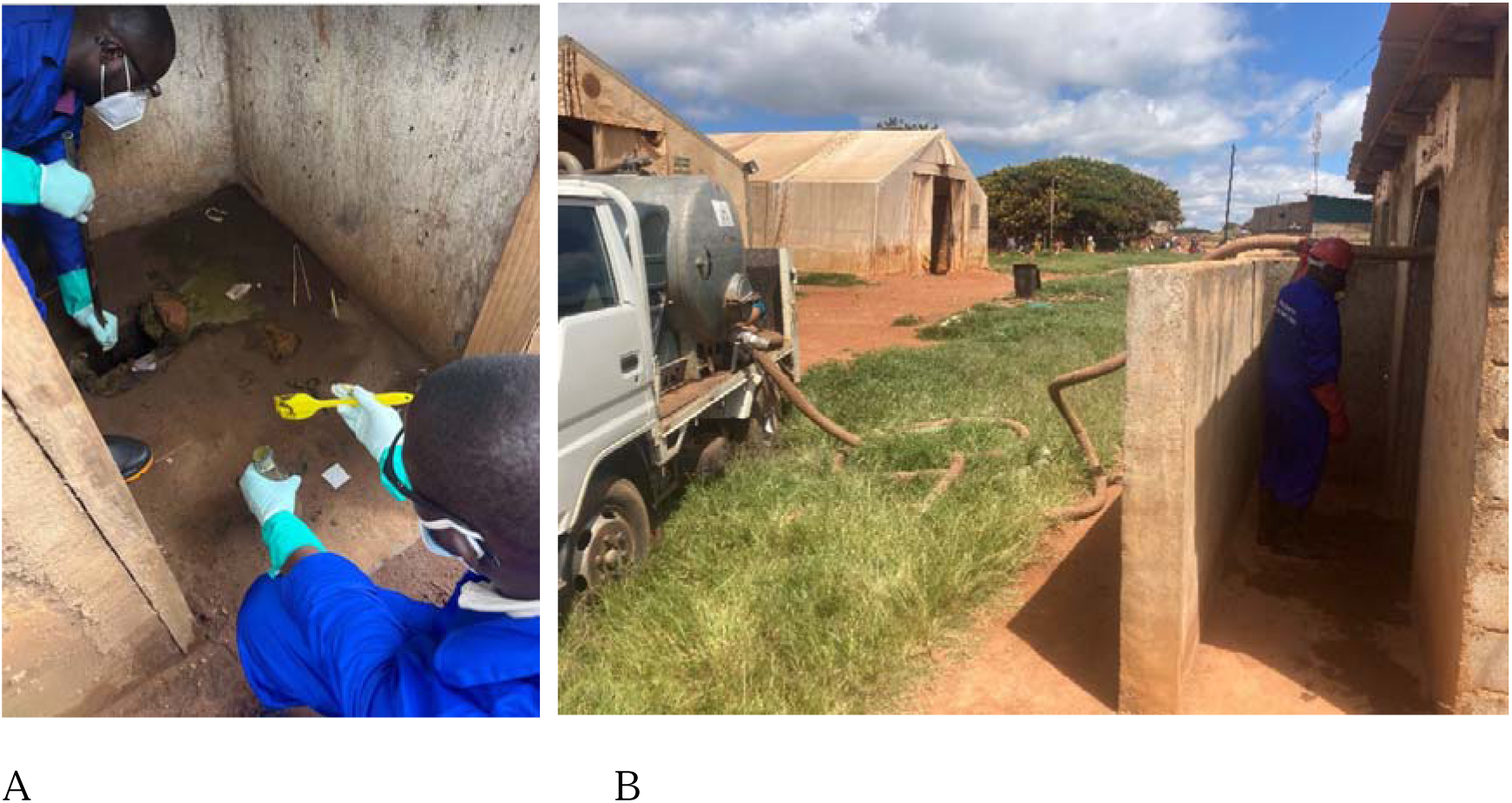
Sample collection, Dzaleka camp. A.) Pit latrine grab sample collection. B.) Vacuum desludging pump truck sample collection.

Pump truck samples were collected by a private contract operator using similar collection procedures. One or two pump-truck samples were collected weekly. These samples were collected after pit latrine grab samples. A commercial pump truck contractor extracted 100– 200 L of fecal sludge from an individual pit latrine into the waste tank and collected the sample directly from the tank using a valve to release the waste directly into the sample bottle (Fig. 2B). The contractor washed the pump truck with water weekly; however, sterilization of the pump truck was not logistically feasible.

### 2.3 Clinical data

The Dzaleka Health Centre provided de-identified data during the study period on the number of cholera vaccinations previously distributed in the camp, the number of diarrhea cases, and confirmed cholera cases through the UNHCR Integrated Refugee Health Information System.

### 2.4 Laboratory analysis

The samples were transported in a cooler box with ice to the Public Health Institute of Malawi (PHIM) National Microbiology Reference Laboratory. The samples were processed using culture methods upon delivery (average time between sample collection and starting the culture, 245 min).

We used our previously-described WES culture method (Chigwechokha et al., 2024). Briefly, clinical methods for culturing *V. cholerae* in stool samples were adapted for WES. For culture, 10 µL of the sample was inoculated on thiosulfate citrate bile salts sucrose agar plates followed by overnight incubation for 18–24 h at 35–37 °C. Presumptive *V. cholerae* colonies were sub-cultured on nutrient agar for another 18–24 h at 35–37 °C. Confirmation analysis (Table S4) was done by either VITEK^®^ MS (*n* = 107) or Analytical Profile Index (API) 20E (*n* = 40) when oxidase was positive. The VITEK^®^ MS quality control used was ATCC 8738. Both confirmation methods were performed according to the manufacturer’s instructions for Enterobacteriaceae and other non-fastidious gram-negative rods, without modifications.

Quality assurance was performed for 15% of the samples (*n* = 22) at the Malawi University of Science and Technology (MUST), which conducts other ongoing WES research (Chigwechokha et al., 2024). Duplicate samples for quality assurance were sent in cooler boxes containing ice packs to MUST, 350 km away from the sampling site, within 24 h of collection, using ground courier services. None of the laboratories was made aware of the others’ results until the weekly stakeholder results-sharing meeting. MUST uses the same techniques for *V. cholerae* culture as PHIM (Chigwechokha et al., 2024), but only uses API for confirmation analysis. Data were manually entered into a password-protected Excel file and shared via email with stakeholders with non-disclosure agreements before weekly data-sharing meetings.

### 2.5 Ethics

This study was reviewed and approved by the Malawi University of Science and Technology Research Ethics Committee (approval no. P.01/2024/117). The University of Louisville Institutional Review Board (23.0969) classified this study as not human research.

## 3. Results and Discussion

This analysis included 147 samples (131 pit latrine and 16 paired pump truck samples) collected over a 19-w period. Twenty-two additional duplicate samples were independently analyzed by MUST for quality assurance. The PHIM culture methods indicated that all (*n* = 147) samples were negative for *V. cholerae* after confirmatory analysis. A duplicate quality assurance sample analyzed by MUST (PT2 on 4/2/2024) was positive after biochemical test confirmation, although the other sample taken from the same latrine on the same day was negative in the PHIM laboratory.

For the samples that had confirmation analysis by VITEK® MS, seven samples showed the presence of other *Vibrio* strains in two or more replicates, including *V. alginolyticus* (*n* = 3), *V. parahaemolyticus* (*n* = 2), and *V. metschikovii* (*n* = 2) (Supplementary Table S5). *Vibrio alginolyticus* and *V. parahaemolyticus* are pathogenic non-cholera *Vibrio spp*. with infections associated with contaminated seafood and direct exposure to water (Baker-Austin et al., 2018). *Vibrio metschikovii* is a less common, non-cholera *Vibrio spp.* (Konechnyi et al., 2021). As these *Vibrio* species have not been well-documented in previous research in Malawi, additional research is needed to understand the transmission routes and interventions to protect camp residents. The Government of Malawi declared that the cholera outbreak officially ended on July 19, 2024 (Republic of Malawi Ministry of Health 2024), coinciding with the end of our WES study. This study builds on previous research on WES for *V. cholerae* in NSSS in Malawi (Capone et al., 2021; Chigwechokha et al. 2024). *Vibrio cholerae* is not a rare target in Malawi; however, we detected it only once in a quality assurance sample using broad surveillance.

The Dzaleka Health Centre reported that 21,640 cholera vaccine doses were administered during the most recent 2021 campaign to camp residents and nearby host community members. The health center reported no confirmed cholera cases during the study period. This indicates a one-dose cholera vaccination coverage of less than 40%, without considering new arrivals or camp attrition from resettlement, repatriation, and death. Although *V. cholerae* was not detected using culture methods beyond one quality assurance sample, cases of diarrhea remained common during the study period. Between February 1, 2024, and July 31, 2024, the Dzaleka Health Centre reported a total of 2,269 cases of diarrhea among Dzaleka residents and Malawian patients (Fig. 3). Among those with diarrhea during the study period, 49% (*n* = 1,108) were children under five years of age. Approximately one-third of the total cases (*n* = 754) involved refugees and asylum seekers, resulting in an average monthly incidence rate of 2.3 cases of diarrhea per 1,000 people. Previous studies have found the incidence of diarrhea in children under five years old living in refugee camps in Africa to be 35.5 cases of diarrhea per 1,000 people per month (Hershey et al., 2011). Further research is required to determine whether this is a result of global reductions in childhood diarrhea morbidity since the initial study by Hershey et al. (2011), a lower incidence of diarrhea in Dzaleka, and/or a reduced tendency to seek public health care in the area.

**Fig. 3.**
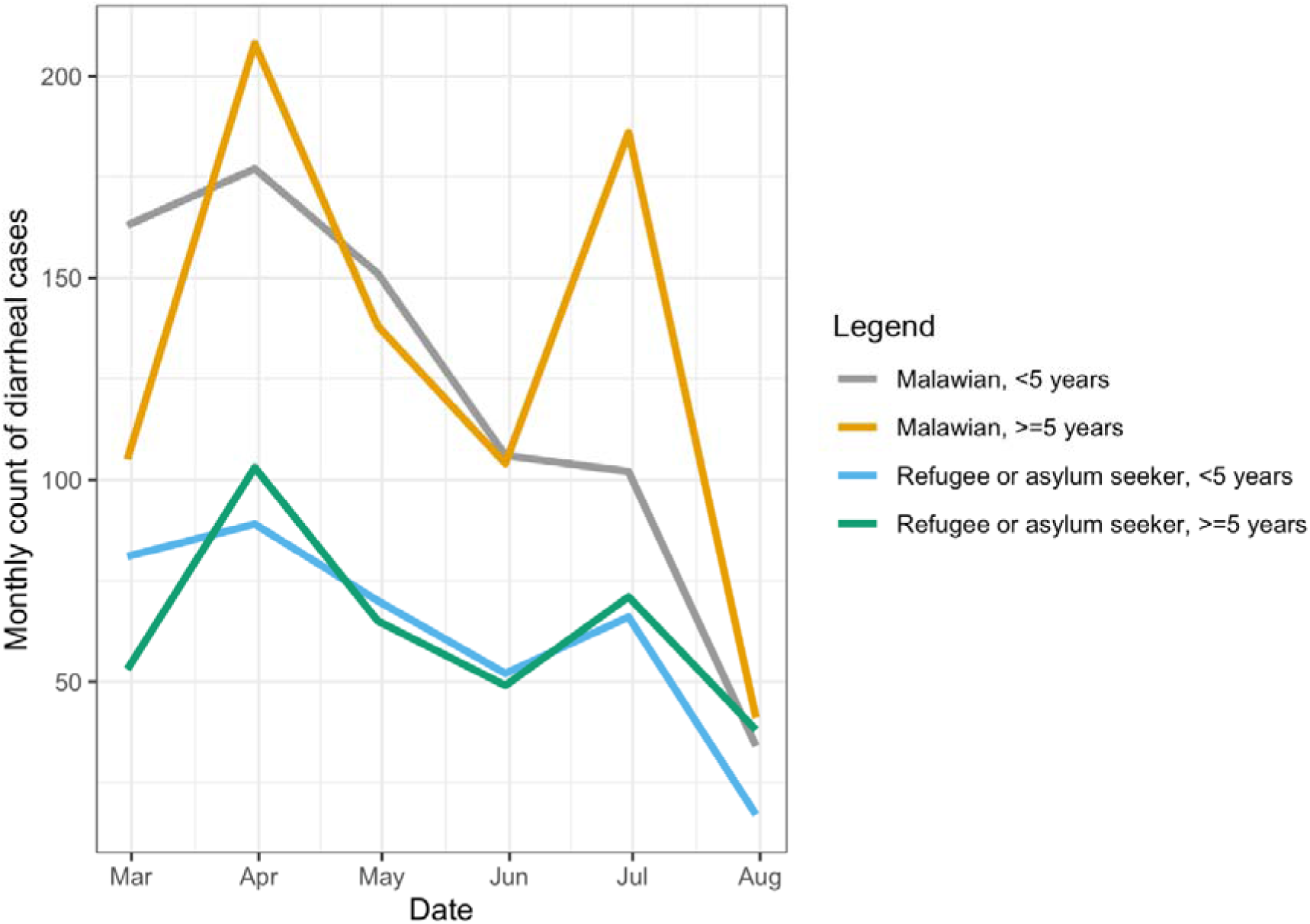
Cases of diarrhea reported by Dzaleka Health Centre during the study period grouped by Malawian and Malawi camp residents, February through July 2024

Pit-latrine user counting indicated that some sampling sites were used more frequently than others (Table S1). The most popular site, PT3, had an average of 10.6 pit latrine users per stance per hour. This indicates that there are more than 100 users per day. The least frequently-used sites, such as PT1 at the reception center and PT4 within the community, were primarily residential. Counting was conducted during the day to ensure researcher safety, and usage may have been differed at these sites in the evening. We estimated that our samples represented deposits from 311 people per sampling day, assuming eight hours of use, or approximately 0.5% of the Dzaleka population. Additional research is needed considering the ethics and minimum sample size for samples to be anonymous from the NSSS when working in a camp.

Pump truck samples were easier to collect than those taken directly from pit latrines, making them a potentially effective WES approach for camps with established desludging operations. Most pump trucks only have a capacity for one cubic meter of fecal sludge, but single latrines often hold a larger volume. Therefore, the pump truck samples are likely limited in representing a single-pit latrine. Further work is needed to determine feasible methods for sterilizing pump trucks between samples or for sampling further down the waste stream, where samples are pooled prior to fecal sludge treatment and/or disposal.

Better and more efficient methods for gathering and reviewing public health data in stretched refugee camps are required. For example, the current WHO WES guidelines (2023) for severe acute respiratory syndrome coronavirus 2 may not be easily extended to WES in a humanitarian context based on circulating pathogens with outbreak variability and NSSS sampling locations, and there is no guarantee of adequate healthcare to detect and treat associated diseases. Many methods used in global WES rely on polymerase chain reaction-based testing, which requires laboratory supplies and capacity that are expensive and difficult to obtain in humanitarian settings. Residents in our study strongly supported the use of WES for communicable diseases such as cholera (Jeboda et al., 2024). The advantage of our culture method approach is that it provides close-to-real-time availability of data; however, the targets that can be measured using this approach are limited. Laboratory partners shared data weekly, approximately four business days after sample collection.

The implications of a positive sample and what to do in response are critically important when considering this type of work. In the event of a positive sample, prior to the start of sample collection, stakeholders agreed to be ready for a range of interventions, including increasing resident access to bucket chlorination (point-of-use treatment), providing supplies for cleaning pit latrines (disinfecting floors), and advocating additional vaccination campaigns to be prioritized. Increasing the water supply was not feasible owing to groundwater limitations in the camp area. The results on WES implementation in Dzaleka will support a wider scale-up in camps globally (Fig. 4).

**Fig. 4.**
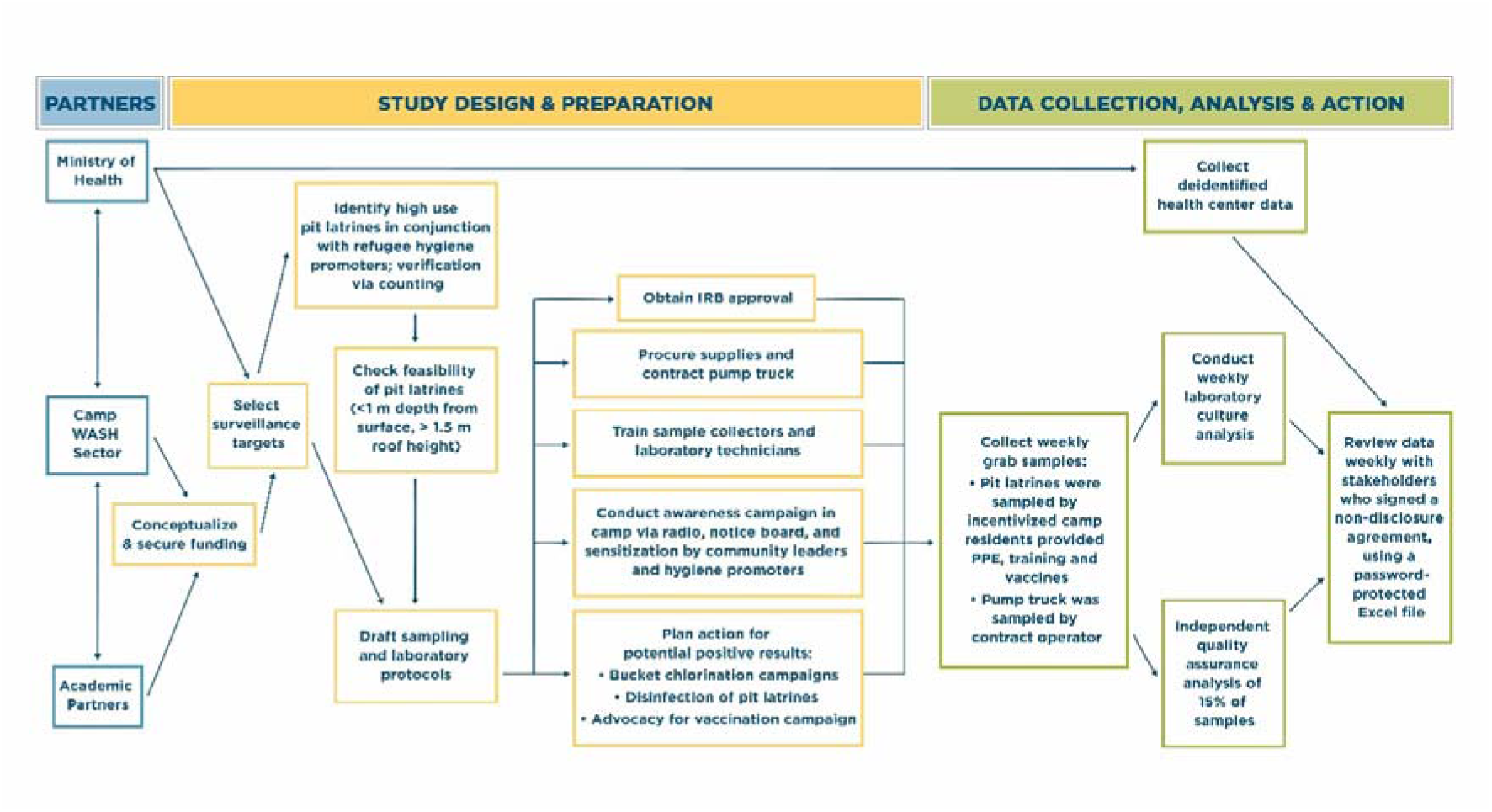
Operational framework for sampling non-sewered sanitation system surveillance at Dzaleka camp including partners, processes, and tools for implementation in a camp setting, Malawi. Project partners included camp WASH cluster, community members, local academics, and the Ministry of Health.

Practical suggestions for increasing the utility of WES among environmental public health practitioners and researchers are critical. The suggested guidelines are:

### Preparatory actions

- Given the declining scope of humanitarian funds, WES may not be feasible without dedicated funding. Funding earmarked specifically for conducting WES from funders with innovation and/or research agendas should be sought.
- The political sensitivity of refugee health data should be considered and mechanisms must be in place to protect data (e.g., non-disclosure agreements and password-protected data) before starting the project.
- Pathogens that, if detected, will change current public health and WASH actions and not just increase knowledge or predetermine subsequent intervention actions in the event of a positive sample should be selected.
- Relationships with local academic and/or government partners with experience in WES and methods that provide technical expertise for applying for institutional review board approval, developing sampling and laboratory protocols, procuring supplies, and conducting research team training should be fostered.
- Refugees and asylum seekers should be engaged to identify the most frequently used public latrines to increase the anonymity and representativeness of the samples.
- Local welders should be approached to manufacture a sampling device.

### During sampling and analysis

- Engage incentivized refugees for sample collection, ensuring adequate protection through access to vaccination and PPE.
- The research team should receive training on donning and doffing PPE, maintaining sample integrity, and site cleanup after collection.
- Report back plans should include weekly data reviews from laboratory partners, camp management, public health partners, and WASH partners.
- The minimum metadata should include site identifiers, sample collection date/time, sample delivery date/time, culture, and confirmation results in a password-protected Excel file.

#### Limitations

The samples in this study were collected at one point in time and space and may not be representative of the entire camp; we were limited by the fact that only certain areas of the camp had high-use pit latrines. Composite samplers were not accessible to the research team and were not logistically feasible for use in pit latrines. It was not feasible to sterilize the desludging truck weekly. In future work, decontamination of the sampling device should consider an ethanol rinse between the sample sites. Further, culture methods via clinical surveillance are less sensitive in detecting *V. cholerae* in stool than PCR (Guillaume et al., 2023).

## 4. Conclusion

*Vibrio cholerae* is a good pathogen candidate for WES in camp settings as multiple WASH preventive interventions are feasible across a range of budgets and timelines by local partners, and some interventions could have been rapidly deployed. The high-use pit latrines and a pump truck offer feasible sample collection in a NSSS camp setting. Although the culture samples were largely negative for *V. cholerae* with the exception of one quality assurance sample, some non-cholerae *Vibrio spp*. were also identified. This project provides a framework for governments, academics, civil society organizations, and refugees to enhance the national public health system through WES. Quantifying pit-latrine usage patterns and catchment areas in relation to health outcomes when conducting WES in camp settings without formal sewer networks is important to ensure that sites are used by enough people and yet remain anonymous. This work provides a significant additional understanding of WES sample design in camp settings to improve human health outcomes and provides a framework for scaling WES efforts in other humanitarian contexts.

## Data Availability

The authors do not have permission to share data.

## Acknowledgements

We are grateful for the expertise provided by David Berendes, Jennifer Murphy, Tom Handzel, Ryan Schweitzer, Matthew Kagoli and Joseph B. Bangoh as well as logistical support from Ernest Chilalika and S. M. Hasanuzzaman. We also thank the sampling team for their assistance with this study.

## Supplementary Material

**Table S1.**
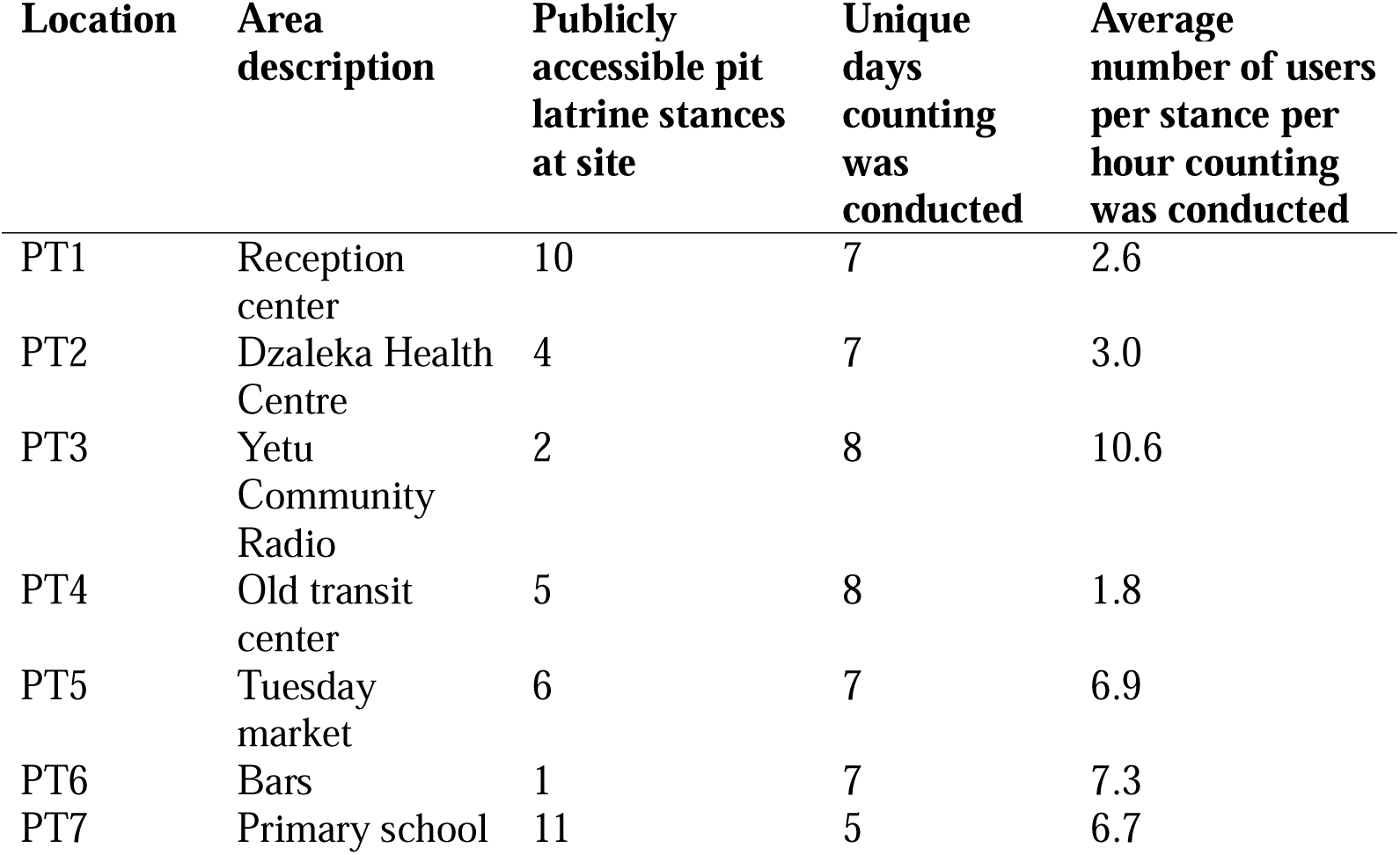
Number of public latrine users at sample sites in Dzaleka camp. Forty-nine latrine counting periods were conducted. The site with the highest use per stance was Yetu Community Radio, with a mean of 10.6 latrine users per hour. The site with the lowest use per stance was the Old transit center, with a mean of 1.8 users per hour. Across sampling sites, there was an average of 5.6 users per stance, per hour of counting.

| **Location** | **Area description** | **Publicly accessible pit latrine stances at site** | **Unique days counting was conducted** | **Average number of users per stance per hour counting was conducted** |
| --- | --- | --- | --- | --- |
| PT1 | Reception center | 10 | 7 | 2.6 |
| PT2 | Dzaleka Health Centre | 4 | 7 | 3.0 |
| PT3 | Yetu Community Radio | 2 | 8 | 10.6 |
| PT4 | Old transit center | 5 | 8 | 1.8 |
| PT5 | Tuesday market | 6 | 7 | 6.9 |
| PT6 | Bars | 1 | 7 | 7.3 |
| PT7 | Primary school | 11 | 5 | 6.7 |

**Table S2.** Description of pit latrine sampling sites (n = 7), Dzaleka camp, Malawi.

| **Location** | **Area description** | **Publicly accessible latrine stances at site** | **Closest water source <sup>a</sup>** | **Site also frequented by members of hosting communities** |
| --- | --- | --- | --- | --- |
| PT1 | Reception center | 10 | Tap water within 20 meters of latrines <sup>b</sup> | No |
| PT2 | Dzaleka Health Centre | 4 | Unchlorinated tap water within 70 meters of latrines | Yes |
| PT3 | Yetu Community Radio | 2 | Unchlorinated tap water within 10 meters of latrines | No |
| PT4 | Old transit center | 5 | Handpump borehole within 80 meters of latrines | No |
| PT5 | Tuesday market | 6 | Handpump borehole within 200 meters of latrines | Yes |
| PT6 | Bars | 1 | Handpump borehole within 100 meters of latrines | Yes |
| PT7 | Primary school | 11 | Handpump borehole within 50 meters of latrines | Yes |

<sup>a</sup>Dzaleka Camp has handpump and motorized boreholes. All water tap water in Dzaleka camp is provided intermittently for a few hours per day due to water rationing.
<sup>b</sup>The source of the water supply for the reception center was changed during the study. In March 2024 it was switched to a chlorinated source.

**Table S3.**
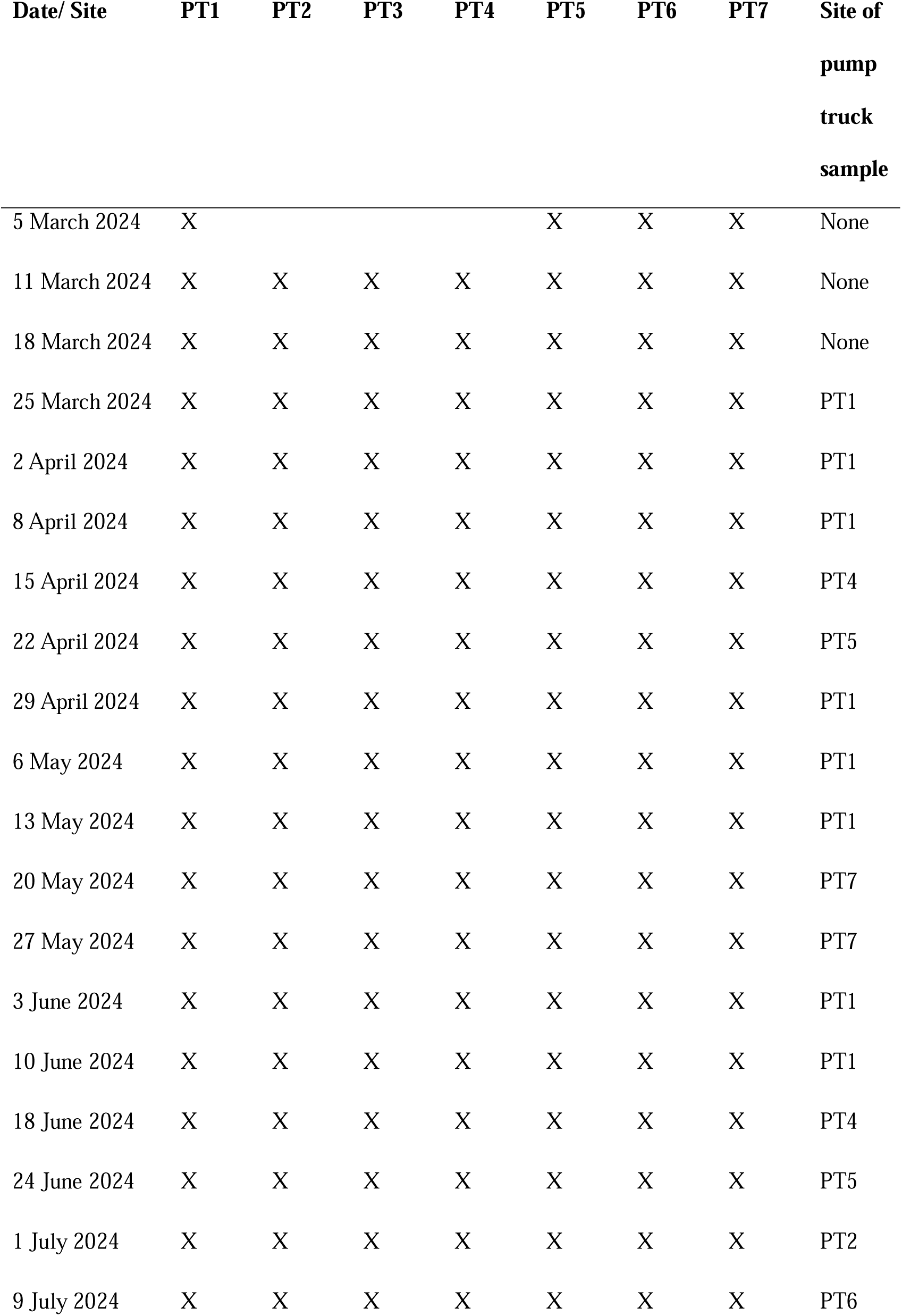
Sampling and analysis details by week for Dzaleka wastewater surveillance for *Vibrio cholerae*.

| **Date/ Site** | **PT1** | **PT2** | **PT3** | **PT4** | **PT5** | **PT6** | **PT7** | **Site of**<br>**pump**<br>**truck**<br>**sample** |
| --- | --- | --- | --- | --- | --- | --- | --- | --- |
| 5 March 2024 | X |  |  |  | X | X | X | None |
| 11 March 2024 | X | X | X | X | X | X | X | None |
| 18 March 2024 | X | X | X | X | X | X | X | None |
| 25 March 2024 | X | X | X | X | X | X | X | PT1 |
| 2 April 2024 | X | X | X | X | X | X | X | PT1 |
| 8 April 2024 | X | X | X | X | X | X | X | PT1 |
| 15 April 2024 | X | X | X | X | X | X | X | PT4 |
| 22 April 2024 | X | X | X | X | X | X | X | PT5 |
| 29 April 2024 | X | X | X | X | X | X | X | PT1 |
| 6 May 2024 | X | X | X | X | X | X | X | PT1 |
| 13 May 2024 | X | X | X | X | X | X | X | PT1 |
| 20 May 2024 | X | X | X | X | X | X | X | PT7 |
| 27 May 2024 | X | X | X | X | X | X | X | PT7 |
| 3 June 2024 | X | X | X | X | X | X | X | PT1 |
| 10 June 2024 | X | X | X | X | X | X | X | PT1 |
| 18 June 2024 | X | X | X | X | X | X | X | PT4 |
| 24 June 2024 | X | X | X | X | X | X | X | PT5 |
| 1 July 2024 | X | X | X | X | X | X | X | PT2 |
| 9 July 2024 | X | X | X | X | X | X | X | PT6 |

**Table S4.** Confirmation analysis method by week for Dzaleka wastewater surveillance for *Vibrio cholerae* at the Public Health Institute of Malawi.

| **Day of sample** | **Confirmation analysis method** |
| --- | --- |
| 5 March 2024 | VITEK® MS |
| 11 March 2024 | VITEK® MS |
| 18 March 2024 | VITEK® MS |
| 25 March 2024 | VITEK® MS |
| 2 April 2024 | API 20E |
| 8 April 2024 | VITEK® MS |
| 15 April 2024 | VITEK® MS |
| 22 April 2024 | VITEK® MS |
| 29 April 2024 | VITEK® MS |
| 6 May 2024 | VITEK® MS |
| 13 May 2024 | VITEK® MS |
| 20 May 2024 | VITEK® MS |
| 27 May 2024 | VITEK® MS |
| 3 June 2024 | VITEK® MS |
| 10 June 2024 | VITEK® MS |
| 18 June 2024 | API 20E |
| 24 June 2024 | API 20E |
| 1 July 2024 | API 20E |
| 9 July 2024 | API 20E |

**Table S5.**
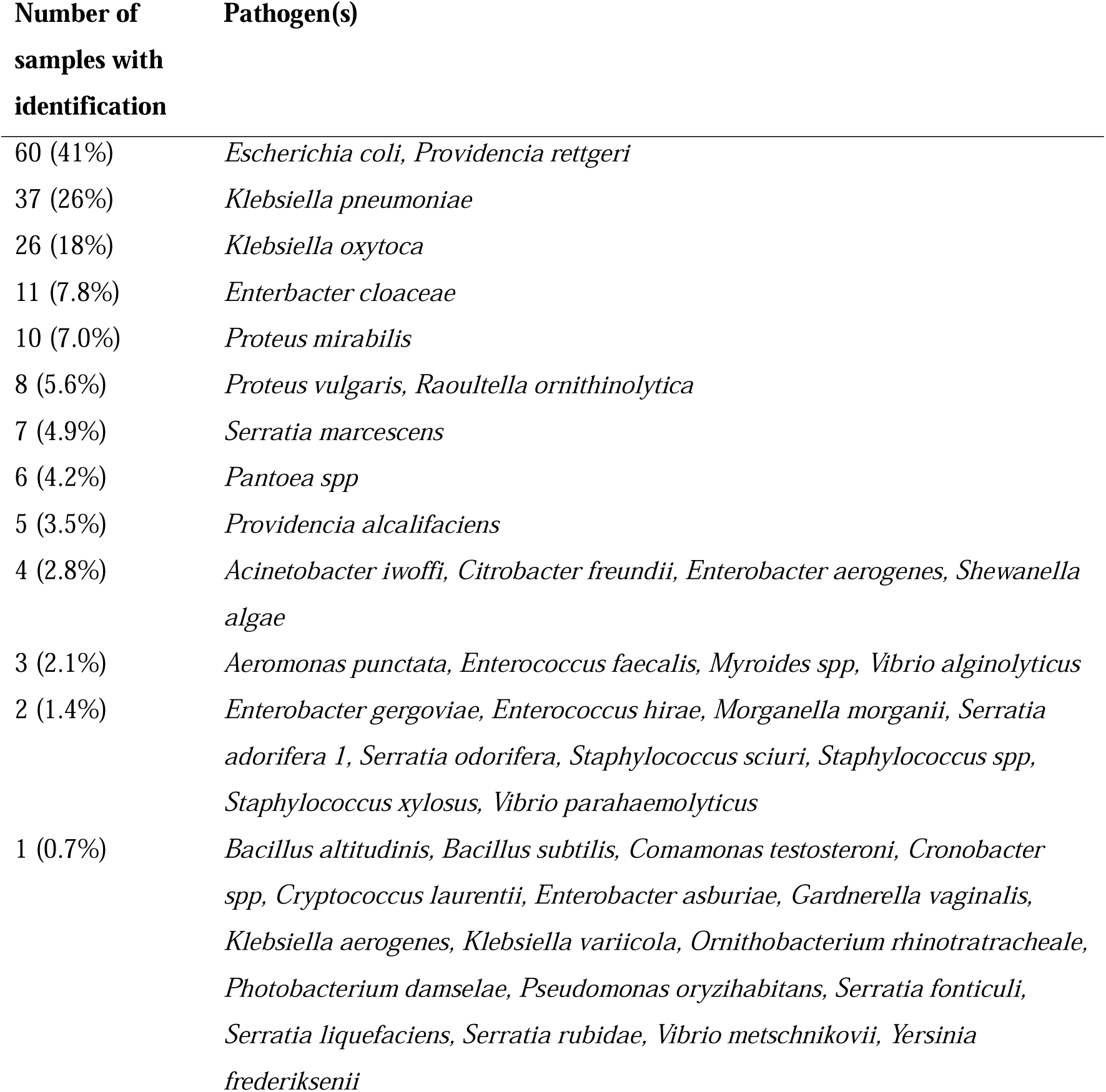
Results of VITEK® MS pathogen identification (*n* = 107) from analysis conducted at the Public Health Institute of Malawi that had full agreement among replicates.

